# Variation of SARS-CoV-2 viral loads by sample type, disease severity and time: a systematic review

**DOI:** 10.1101/2020.09.16.20195982

**Authors:** Thomas Edwards, Victor S. Santos, Anne L. Wilson, Ana I. Cubas-Atienzar, Konstantina Kontogianni, Christopher T. Williams, Emily R. Adams, Luis E. Cuevas

## Abstract

**Background:** To describe whether SARS-CoV-2 viral loads (VLs) and cycle thresholds (CTs) vary by sample type, disease severity and symptoms duration.

**Methods:** Systematic searches were conducted in MEDLINE, EMBASE, BioRxiv and MedRxiv. Studies reporting individual SARS-CoV-2 VLs and/or CT values from biological samples. Paired reviewers independently screened potentially eligible articles. CT values and VLs distributions were described by sample type, disease severity and time from symptom onset. Differences between groups were examined using Kruskal-Wallis and Dunn ‘s tests (post-hoc test). The risk of bias was assessed using the Joanna Briggs Critical Appraisal Tools.

**Results:** 14 studies reported CT values, 8 VLs and 2 CTs and VLs, resulting in 432 VL and 873 CT data points. VLs were higher in saliva and sputum (medians 4.7×108 and 6.5×104 genomes per ml, respectively) than in nasopharyngeal and oropharyngeal swabs (medians 1.7×102 and 4.8×103). Combined naso/oropharyngeal swabs had lower CT values (i.e. higher VLs) than single site samples (p=<0.0001). CT values were also lower in asymptomatic individuals and patients with severe COVID-19 (median CT 30 for both) than among patients with moderate and mild symptoms (31.4 and 31.3, respectively). Stool samples were reported positive for a longer period than other specimens.

**Conclusion:** VLs are higher in saliva and sputum and in individuals who are asymptomatic of with severe COVID-19. Diagnostic testing strategies should consider that VLs vary by sample type, disease severity and time since symptoms onset.

**Summary:** This systematic review found a higher viral load in saliva and sputum than in nasopharyngeal swabs, in asymptomatic individuals and patients with severe COVID-19. Diagnostic testing strategies should consider the type of sample, disease severity and the time since symptoms onset.

## INTRODUCTION

In late 2019, the emergence of Severe Acute Respiratory Syndrome Coronavirus 2 (SARS-CoV-2) triggered the Coronavirus Disease 19 (COVID-19) pandemic [1,2], which has now affected almost all countries, with devastating consequences. Even though most infections remain asymptomatic or have mild presentation [1–4], a significant proportion progress to severe COVID-19 with high case fatality rates and over 600,000 deaths worldwide by July 2020.

The detection of SARS-CoV-2 infections relies on amplifying the viral ribonucleic acid (RNA) using reverse transcription quantitative polymerase chain reaction (RT-qPCR) assays; multiple “in-house” and commercial assays are available [5], with assay sensitivity varying from 1 to 100 viral genome copies per reaction. The amount of virus detected is thought to vary according to the specimen examined, the time of sampling after symptoms onset and disease severity, although surprisingly there are no systematic reviews describing these variations [6–9]. There is conflicting information on the association of VLs and disease severity. In some studies high VLs are associated with severe COVID-19, as an indication of higher viral replication, cellular damage and pathology[10], which trigger multiorgan failure [11]; however, not all studies have found this association and asymptomatic individuals can have high VLs [12]. Saliva and sputum tend to have higher VLs [13,14], although nasal and oropharyngeal samples are used more frequently [15–17]. Virions can be detected two days before symptom onset when VLs are high, and generally remain detectable for 7-21 days. However, some patients have detectable RNA for up to two months and virus excretion is more prolonged in stools [18,19].

Most RT-qPCR assays report cycle threshold (CT) values or estimate the viral loads (VL) utilising standard curves, which are assay specific [20]. Most reports therefore include either the CT or the VL, as extrapolation from CT values [21], but rarely both values.

Given the heterogeneity of findings, we conducted a systematic review of SARS-CoV-2 CT values and VLs reported in the literature. Herein we describe virus excretion kinetics and how these titers differ with disease severity, time from symptom onset, type of sample tested and the RT-qPCR assay used.

## METHODS

### Search strategy and selection criteria

The primary questions were whether VLs and CT values varied according to sample type, disease severity (asymptomatic, mild and severe), the time since the start of symptoms and RT-qPCR assay. The search was conducted within the University of Bern Living Evidence on COVID-19 database (https://ispmbern.github.io/covid-19/living-review/), which uses the following search terms: MEDLINE: (“Wuhan coronavirus” [Supplementary Concept] OR “COVID-19” OR “2019 ncov”[tiab] OR ((“novel coronavirus”[tiab] OR “new coronavirus”[tiab]) AND (wuhan[tiab] OR 2019[tiab])) OR 2019-nCoV[All Fields] OR (wuhan[tiab] AND coronavirus[tiab]))))) EMBASE: ncov OR (wuhan AND corona) OR COVID-19 and BioRxiv/MedRxiv: ncov or corona or wuhan or COVID. The database was downloaded with a cut-off data of 8^th^ April 2020. We sub searched the database by retaining literature that contained “load” OR “dynamic*” OR “diagnostic*” OR “shed*” OR “serial” in the title or abstract. We also searched the bibliography of systematic reviews and modelling studies to identify additional studies. No language restrictions were applied, and we assessed the English abstracts of Chinese language manuscripts for inclusion and exclusion criteria. Two reviewers (ALW and CTW) independently conducted screening on the title, abstract and, if required, the full text of the manuscript. We included preprints and a second reviewer (VSS) verified all articles excluded during the full-text search. Studies were retained if they had used RT-qPCR and had reported VLs or CT values of SARS-CoV-2 in respiratory samples (oropharyngeal, nasopharyngeal, saliva, sputum, endotracheal aspirate or bronchoalveolar lavage) or stools, at one or more timepoints from individual patients in the community, outpatient or inpatient settings. Studies were included if they reported individual patient data or graphical displays that allowed data extraction and if it was possible to ascertain the day of sampling in relation to the initiation of symptoms. We included single case reports, case series and cohort studies. We excluded publications without original data, those which reported aggregated data, when data extraction of individual values was not possible and those with potentially overlapping data with other studies. The study protocol is registered at PROSPERO (Registration number: CRD42020181955).

Data extraction and bias assessment

Data were extracted in duplicate (by TE, VSS, ALW, ACA, KK and CTW) using a pre-piloted extraction form. Disagreements were resolved by discussion and involvement of a third party, where necessary. Graphs were digitised using Engauge software (http://markummitchell.github.io/engauge-digitizer/). Extracted data included author, country, participant characteristics, and clinical setting. Individual patient data was retrieved including disease severity (asymptomatic, mild, moderate, severe), as defined by the study authors, frequency and day of symptoms, sample type, diagnostic assay, CT values and/or VLs. When standard curves for the RT-qPCR assays were available, we converted CT values into viral enome copies per ml to allow their comparison with studies only reporting VLs. Standard curves were available for the Corman 2020 RT-qPCR, the China CDC N and ORF1b RT-qPCR, the United States CDC N1, N2, N3 RT-qPCRs [20] and the BGI Schenzen ORF1ab [22]. The values of converted VLs were included in both the CT values and VLs analyses. Although this method is not as accurate as when the standard curves are run experimentally by the study authors, it allows a good approximation for the conversion of CT values into VLs.

The Joanna Briggs Critical Appraisal Tools was used to assess the risk of bias and study quality for case report and case series, as appropriate [23]. After critical appraisal of each item, the studies were rated as good, fair, or poor and the findings were discussed qualitatively.

### Statistical analysis

All laboratory data were standardised to display similar units and presented as medians and interquartile ranges (IQR). Categorical variables were described using descriptive statistics with 95% confidence intervals (95%CI). The normal distribution of continuous data was verified using the Kolmogorov-Smirnov test. As most CT values and VLs had skewed distributions, we used nonparametric tests to verify the statistical significance of the differences between groups. Kruskal-Wallis tests were used to assess differences between CT values and VLs by disease severity, sample type and time from symptom onset. If Kruskal-Wallis ‘s tests were significant, we performed multiple comparisons using Dunn ‘s tests (post-hoc test) to determinate differences between groups. Lines were fitted to longitudinal data sets using non-linear regression or polynomial models depending on the suitability of the data. P values <5% were considered statistically significant. Data were analysed using GraphPad Prism Version 5 (GraphPad Software, Inc., San Diego, CA).

## RESULTS

The search strategy identified 746 reports. After screening titles and abstracts, 124 full-text articles were assessed for eligibility and 24 studies [1,7,12,14,16,17,24–41] met the inclusion criteria (Supplementary Figure 1). Most studies were rated good (12/24) or fair (9/24) for risk of bias, with three rated poor (Supplementary Tables 1 and 2). Eighteen studies were case series [1,12,14,16,17,26–29,31,32,34,36–41] and six case reports [7,24,25,30,33,35] (Table 1). Most studies included hospitalized patients and only two reported outpatients ‘data. CT values were reported in 18 studies [1,7,12,16,24,26,27,29,30,32–35,37–41,], VLs in six studies [14,17,25,28,31,36] and in six studies [7,12,29,33,35,37] with published standard curves CT values were converted to VLs. Individual patient data was identified for 173 participants yielding a database of 873 CT values and 494 VL data points and their characteristics are shown in Table 2.

**Table 1:** Characteristics of included studies.

| Author | Country | Study design | Setting | Number of patients | Sample type | PCR target gene | CT or VL reported |
| --- | --- | --- | --- | --- | --- | --- | --- |
| Caly L. | Australia | Case report | In-patient | 1 | NP, SP, U, SE, R | RdRP | CT |
| Chan J. | China | Case series (family cluster) | OPD, in-patient | 6 | NP, OP, R, U, SE, P, SP | S | CT |
| Chen D. | China | Case report | In-patient | 1 | OP | Unknown | VL |
| Holshue M.L. | China | Case report | In-patient | 1 | OP | N1, N2, N3 | CT* |
| Kam K. | Singapore | Case series | In-patient | 2 | NP, R, U, B | ORF1ab, N | CT |
| Kim E. | Korea | Case series | In-patient | 28 (9 with viral kinetics) | OPNP, SP | E, RdRP | CT |
| Kim J.Y. | Korea | Case series | In-patient | 2 | OPNP, SP, SE, U, R, P (viral kinetics only available for OPNP and SP) | E | VL |
| Le T. | Vietnam | Case series / cluster analysis | In-patient | 24 (12 with data) | OP | E, RdRP, N | CT* |
| Lee N. | Taiwan | Case report | In-patient | 1 | OP, NP (CT value reported for OP only) | E, RdRP1, N | CT |
| Lescure F. | France | Case series / cohort study | In-patient (3 in ICU) | 5 | NP (aspirate or swab), R, B | E, RdRP | VL |
| Liu Y. | China | Case series | In-patient (several ICU) | 12 | OP, BALF | N, ORF1ab | CT |
| Pan Y. | China | Case series | In-patient | 82 (2 with IPD) | OP, SP, U, R | Unknown | VL |
| Scott, S. | USA | Case report | OPD | 1 | OP, NP, SE, SP | N1, N2, N3 | CT* |
| Shen C. | China | Case series | ICU | 5 | NP | N, ORF1ab | CT |
| Tan L. | China | Case series | In-patient | 2 (viral load on 1 patient) | OP | RdRP/E | CT* |
| Tan L. | Vietnam | Case report | In-patient | 1 | OP, R, P, U | RdRP | CT* |
| To K.K.W. | Hong Kong, | Case series | In-patient | 12 | SA | S | VL |
|  | China |  |  |  |  |  |  |
| Woelfel, R. | Germany | Case series | In-patient | 9 | OP, SP, R | RdRP | VL |
| Xu Y. | China | Case series / screening programme | In-patient | 10 | NP, R | N, ORF1ab | CT* |
| Yang Y. | China | Case series | In-patient | 10 | OP, NP, SP, BALF | N, ORF1ab | CT |
| Young B. | Singapore | Case series | In-patient (2 patients in ICU) | 18 | NP, B, U, R | N, S, ORF1ab | CT |
| Zhang N. | China | Case series | In-patient | 10 | OPNP, R | ORF1ab | CT |
| Zhang W. | China | Case series | In-patient | 13 | OP, R | ORF1ab | CT |
| Zou L. | China | Case series / cluster analysis | Unclear | 17 | OP, NP | ORF1ab | CT |

IPD: individual patient data
OPD: out-patient, ICU: intensive care unit
OP: oropharyngeal, NP: nasopharyngeal, OPNP: combined oropharyngeal and nasopharyngeal, SA: saliva, SE: serum, SP: sputum, U: urine, B: blood, R: rectal/faecal/stool, BALF: Bronchoalveolar Lavage Fluid, P: plasma, B: blood, P: plasma
\*CT values converted to VLs using published standard curves for the RT-qPCR used in the study

**Table 2.** Characteristic of the 173 patients included.

| **Variables** | **N (%)** |
| --- | --- |
| Age, median (IQR*) in years | 46 (28.5-62.5) |
| Sex |  |
| Male | 45 (26.0) |
| Female | 45 (26.0) |
| Not reported | 83 (48.0) |
| Comorbidity, yes | 26 (15.0) |
| Type of comorbidity** |  |
| Hypertension | 11 (30.6) |
| Diabetes | 5 (13.9) |
| Heart disease | 4 (11.1) |
| Smoker | 4 (11.1) |
| Cancer | 3 (8.3) |
| Lung diseases | 2 (5.6) |
| Chronic renal disease | 2 (13.9) |
| Other | 5 (13.9) |
| Symptoms*** |  |
| Fever | 81 (59.5) |
| Cough | 65 (47.8) |
| Diarrhoea | 17 (12.5) |
| Sore throat | 11 (8.1) |
| Shortness of breath | 11 (8.1) |
| Headache | 9 (6.6) |
| Chest pain | 7 (5.1) |
| Myalgia | 6 (4.4) |
| Fatigue | 3 (2.2) |
| Stage of disease |  |
| Asymptomatic | 3 (2.2) |
| Mild | 80 (58.8) |
| Moderate | 2 (1.5) |
| Severe | 33 (24.3) |
| Not reported | 18 (13.2) |
| ARDS | 11 (8.1) |
| Bacterial pneumonia | 11 (11.0) |
| Outcome |  |
| Continued follow up | 35 (25.7) |
| Discharged | 34 (25.0) |
| Death | 1 (0.7) |
| Not reported | 103 (59.5) |

\* IQR: interquartile range
\*\*Some patients had more than one comorbidity.
\*\*\* Some patients had more than 1 symptom.

**Figure 1.**
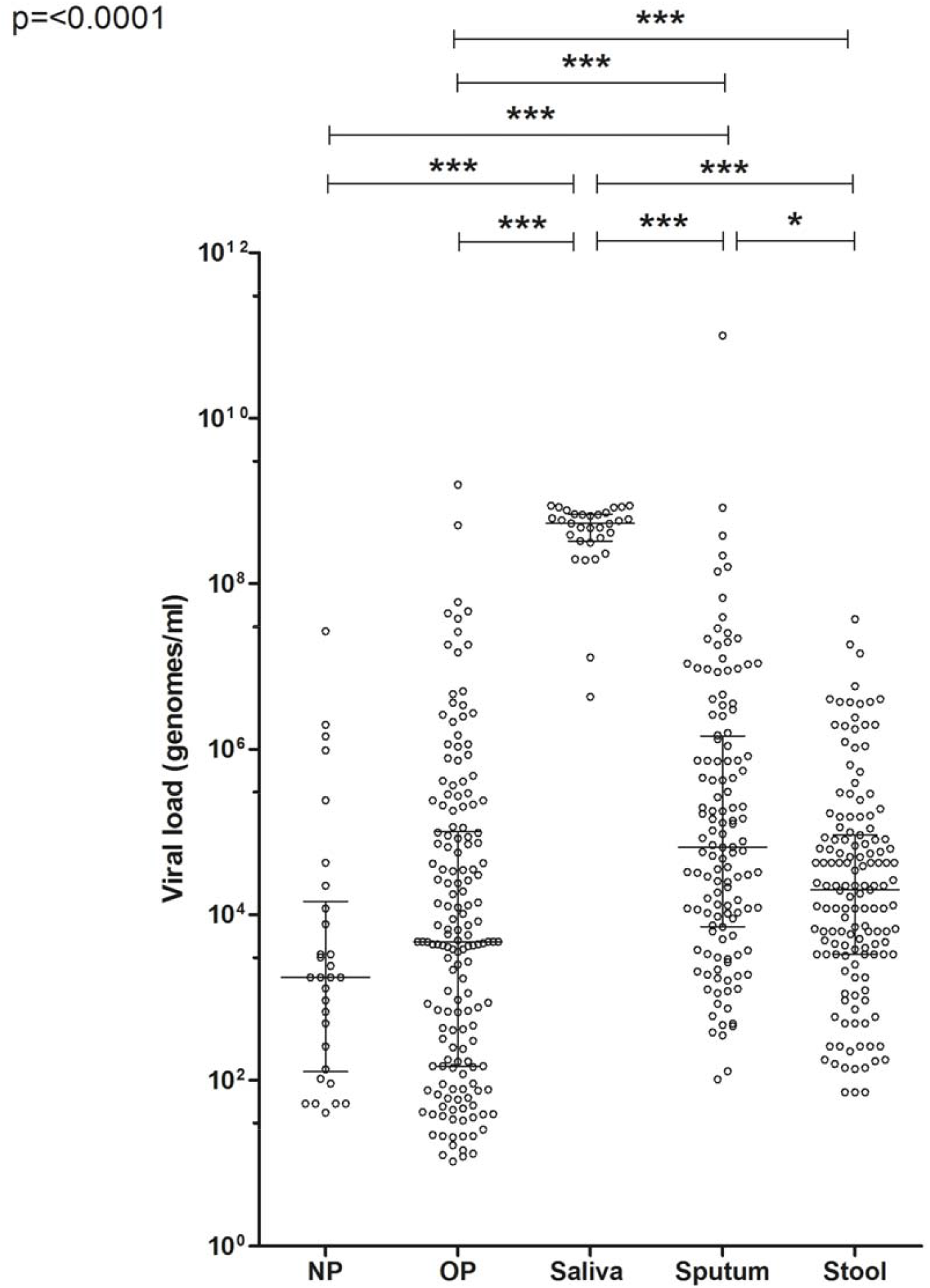
Distribution of viral loads by sample type. Error bars indicate medians and interquartile range (NP = nasopharyngeal, OP = oropharyngeal).

### Viral loads

The median VL across all sample types was 2.3×10^4^ viral genomes per ml and ranged from 12 to 1×10^11^ genomes per ml (Supplementary Figure 2). VLs differed by samples type (P <0.0001) (Figure 1) and were highest in saliva (median 4.7×10^8^ genomes per ml) and sputum (6.5×10^4^ genomes per ml). Nasopharyngeal and oropharyngeal swabs had median VLs of 1.7×10^3^ and 4.8×10^3^ per ml, respectively, which were lower than in saliva (P <0.001) and sputum (P <0.001). VLs declined with the number of days since symptoms onset (Figure 2) and were reported for a longer time period from stool (28 days) than oropharyngeal (22 days) and nasopharyngeal swabs (18 days). The large VL in Saliva samples was maintained for the duration of positivity, however VLs in saliva were not reported beyond 12 days.

**Figure 2.**
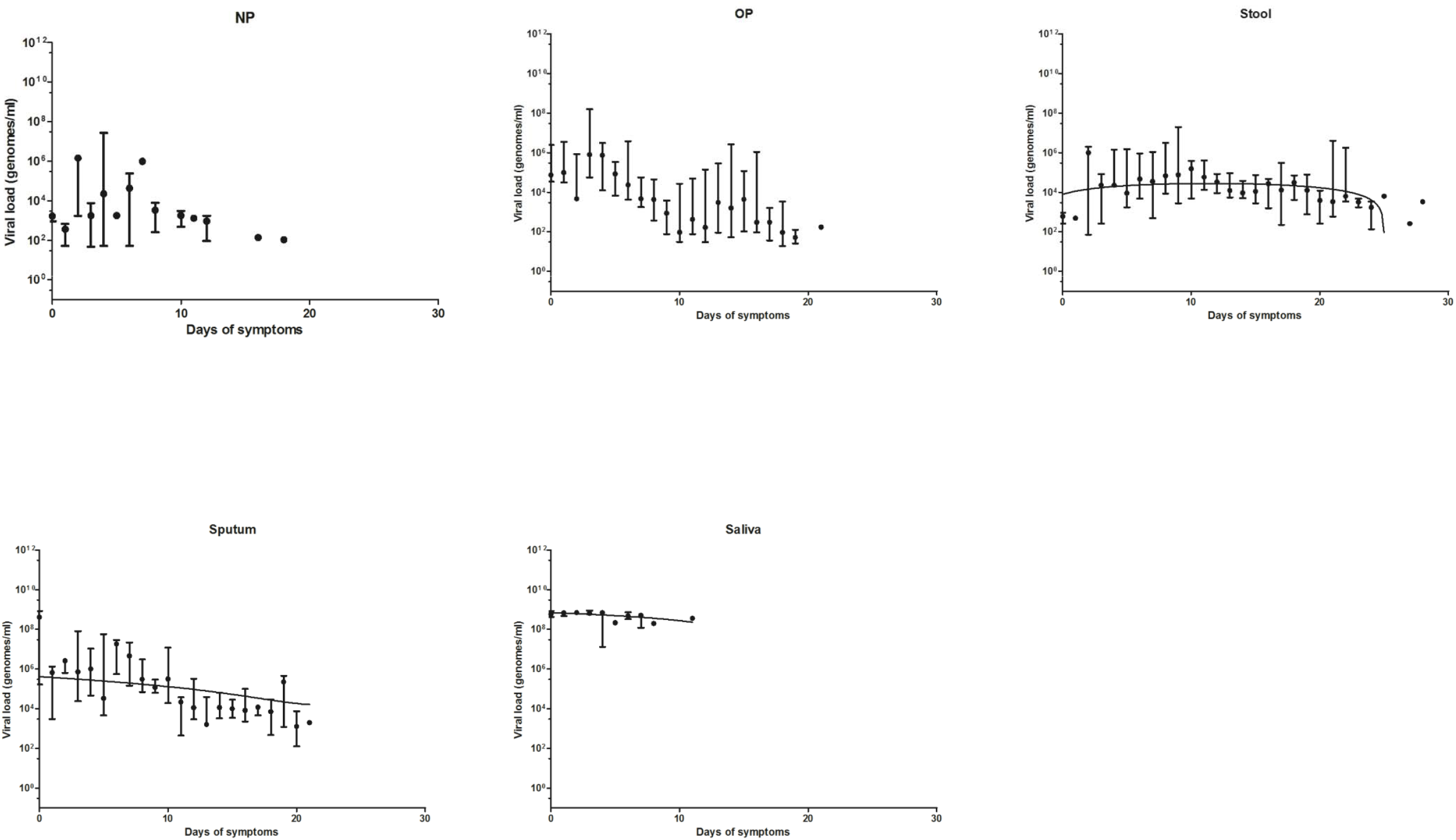
Viral loads reported after symptoms onset by sample type. Error bars indicate medians and interquartile range (NP = nasopharyngeal, OP = oropharyngeal).

CT values The median CT values across all samples was 31.2, and ranged from 14.3 to 39.6, (Supplementary Figure 3). Values differed slightly with disease severity (P = 0.0119, Figure 3), as severe COVID-19 cases had median CT values of 30, compared with 31.4 and 31.3 for moderate and mild cases. However asymptomatic individuals also had a median of 30, which was not statistically different from severe COVID-19 cases (P >0.05). CT values also differed by sample type, with median CTs of 32.2 for nasopharyngeal, 31.9 for oropharyngeal, 31.7 for stools and 27.3 for sputum (P <0.0001, Figure 4). Combined naso/oropharyngeal swabs had lower median CTs (29.5) than other samples over the first 10 days of symptoms. Median CT values increased over time (indicating lower VLs) (Figure 5) and positive samples were reported for longer periods from nasopharyngeal swabs than oropharyngeal swabs.

**Figure 3.**
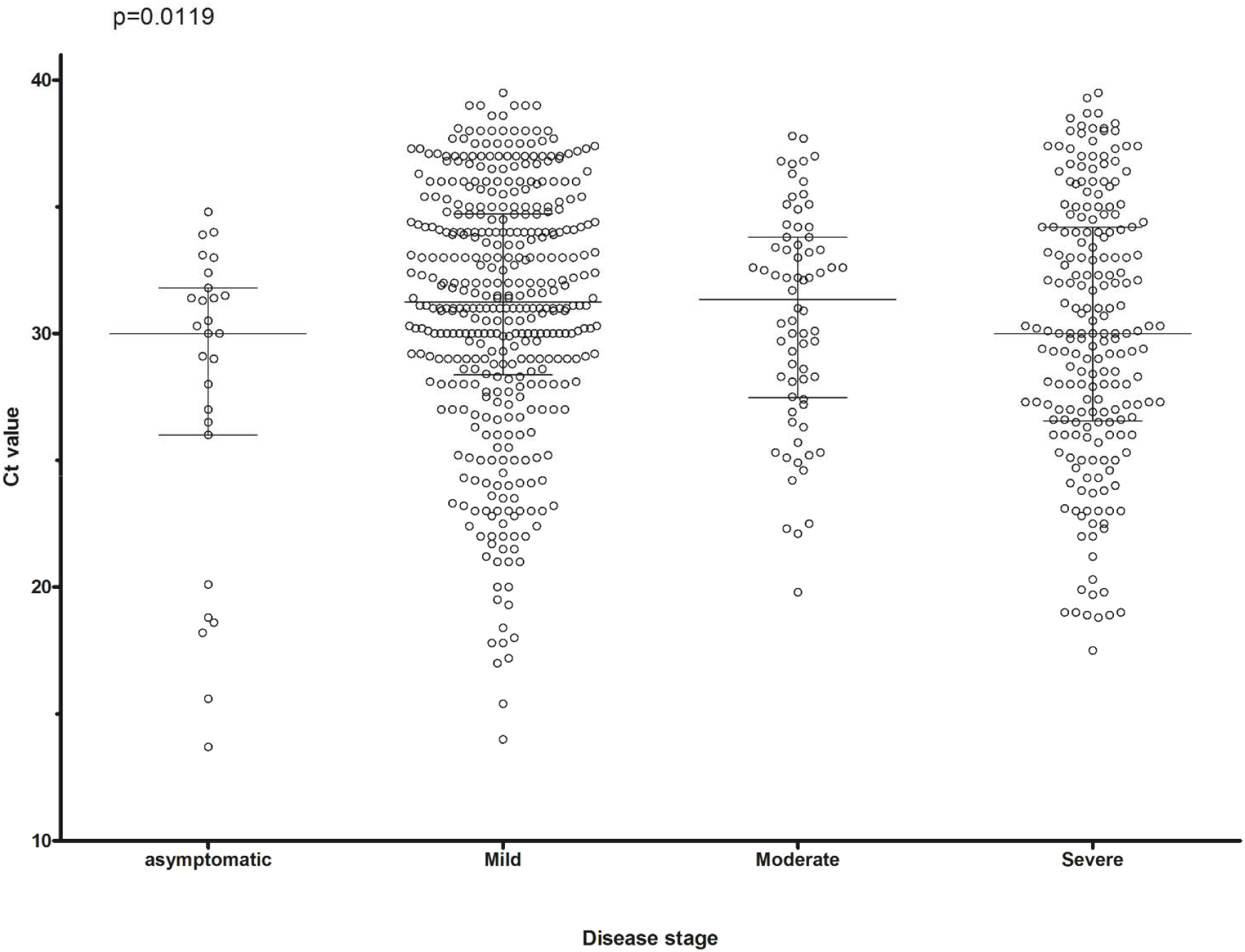
CT values by disease severity. Error bars indicate medians and interquartile range.

**Figure 4.**
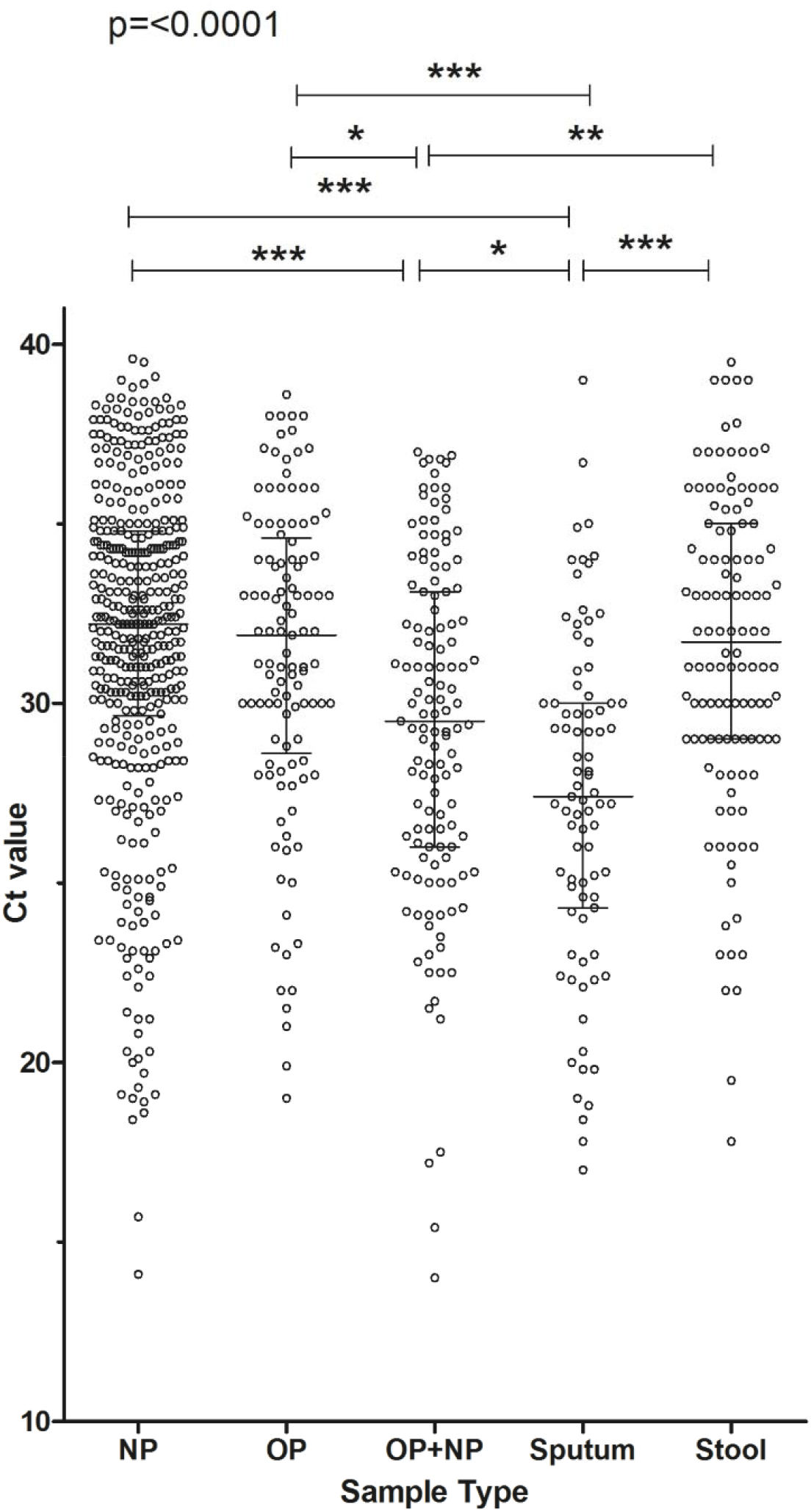
CT values by sample type. Error bars indicate medians and interquartile range (NP = nasopharyngeal, OP = oropharyngeal).

**Figure 5.**
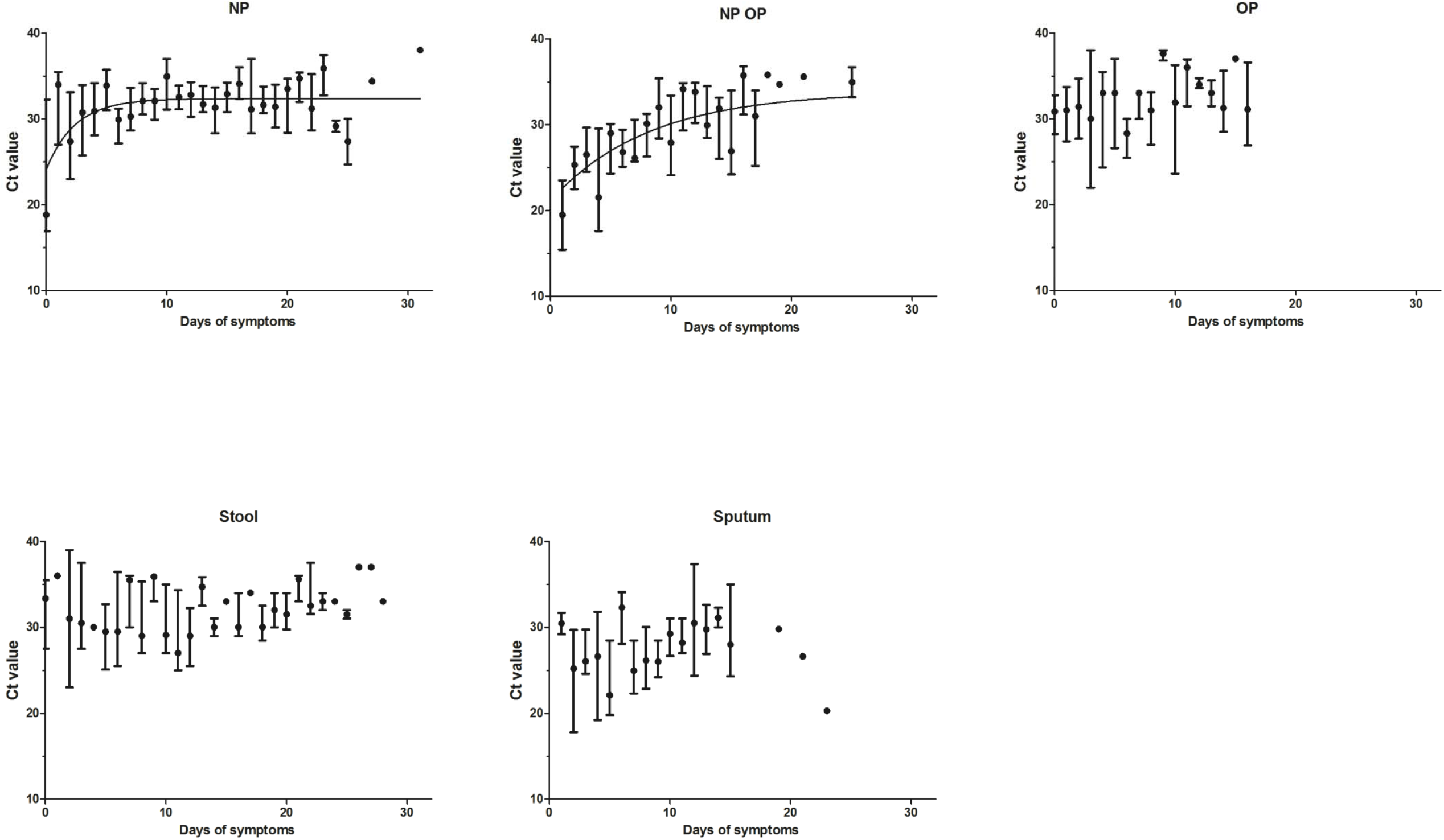
CT values reported after symptoms onset by sample type. Error bars indicate medians and interquartile range (NP = nasopharyngeal, OP = oropharyngeal).

CT values from nasopharyngeal swabs varied with disease severity. Severe cases continued to have positive results recorded for up to 31 days, while the last reported positive result for mild cases was 21 days (Supplementary Figure 4). Finally, CT values varied with the target gene of the RT-qPCR assays (Supplementary Figure 5), with the E gene target providing the lowest CT values, which were statistically significantly lower than the CT values reported for Orf1ab and RdRp (P <0.0001).

## DISCUSSION

CTs and VL titers varied by sample type, with saliva and sputum having higher VLs (and lower CTs), than other respiratory samples. Saliva and sputum are easier to collect and less invasive than nasopharyngeal swabs, especially among patients with mild and moderate COVID-19, and can be sampled by the patient, reducing the risk of infection to healthcare workers. However, all saliva datapoints were obtained from a single study [36], and a follow on publication after the search cut-off date of 39 SARS-CoV-2 positive patients suggested a lower VL in comparison with nasopharyngeal samples [42]. Sputum, which is a diagnostic sample used for other respiratory conditions, had higher VLs and detectable virions for a longer time than other respiratory samples. This may be attributed to RNA from the lungs taking longer to be excreted from the lower respiratory tract, possibly by mechanical trapping in necrotic tissues or to lower airway cells containing greater density of human angiotensin converting enzyme 2 receptors, which are the target for virus binding [43]. VLs in the oropharynx were slightly higher than in the nasopharynx, although this was not statistically significant, as reported in studies not included here [44]. However, combining samples from both sites into a single sample resulted in higher VLs (lower CT values), which has been reported for other viruses [45]. This may reflect that the virus is not always detected simultaneously from all anatomical sites, and multiple-site swabs can decrease false negative results by extending the sampling area. Independently of the underlaying reasons, sampling both sites seem to increase the yield for upper respiratory tract specimens.

Our study highlights the presence of high VLs in stools, with shedding recorded up to 28 days post symptoms onset. Although RT-qPCR does not distinguish whole viruses from RNA fractions [15], stool examination is gaining public health importance for surveillance, as its recovery from sewage samples seems to loosely correlate with the levels of infection in the community [46] This also has the potential to forewarn of epidemics [47].

Although VLs have been considered markers of COVID-19 severity[48], with potential as prognostic biomarkers [47], there is no consensus across studies [49,50]. In our analysis, CT values were slightly lower (higher VLs) in patients with asymptomatic infections and severe COVID-19 than among patients with mild and moderate presentations. Moreover, CT values had wide ranges with significant overlap across the groups, limiting their value as prognostic markers.

RT-qPCR assays targeting the E gene generally reported lower CTs than other targets. This is potentially due to many studies using the Charité RT-qPCR assays [5], of which the E gene primer/probe set has been shown to be more sensitive than the RdRp set [51]. The lower CTs obtained with the E gene may not be indicative of inherent increased sensitivity with this PCR target, but demonstrate the high sensitivity of this RT-qPCR assay.

Our data has several limitations that need to be considered to interpret the data. Most studies were case reports or case series and therefore testing was initiated by clinical need or patient management. It is thus possible that data was collected from patients with unusual conditions and does not represent the whole spectrum of disease. Data also represents early reports during the pandemic, when diagnostic capacity was limited, and testing was prioritised for patients with the greatest need. Therefore, VLs may have over-represented patients with symptomatic infections and their contacts and further studies are needed to further document VLs in asymptomatic and mild cases through community surveillance. Other potential limitations include the use of the disease severity classifications of the authors, which were not standardised until later in the epidemic. Moreover, we included pre-prints that had not been peer-reviewed, with potentially varying quality. Despite these limitations, few studies had a significant risk of bias and the main issues noted were data aggregation and a lack of timelines to describe the patient clinical progression.

In conclusion, we have demonstrated that SARS-CoV-2 CTs and VLs vary between sample type, time point, and disease severity. This information will be useful for the selection of specimens for SARS-CoV-2 confirmation, the development of new diagnostic assays and a better understanding of the patterns of VLs over time to be expected among the different specimens. This information will also be useful to inform sample pooling strategies, in which samples from several patients are ‘pooled ‘and tested together, which are being explored as a resource saving approach to increase diagnostic capacity and for large scale screening of asymptomatic populations.

## Data Availability

Data is available from the corresponding author by request

